# Features of C-reactive protein in COVID-19 patients with different ages, clinical types and outcomes: a cohort study

**DOI:** 10.1101/2020.10.26.20220160

**Authors:** Gaojing Qu, Guoxin Huang, Meiling Zhang, Hui Yu, Xiaoming Song, Haoming Zhu, Lei Chen, Yunfu Wang, Bin Pei

**Author notes:** Address correspondence to author Bin Pei at: Evidence-Based Medicine Center, Xiangyang No.1 People’s Hospital, Hubei University of Medicine, 15 Jiefang Road, Fancheng District, Xiangyang 441000, Hubei, China. And address correspondence to author Yunfu Wang at: Department of Neurology, Taihe Hospital, Hubei University of Medicine, 30 South Renmin Road, Shiyan, Hubei 442000 China. Gaojing Qu and Guoxin Huang contributed equally and should be considered first authors. Bin Pei and Yunfu Wang contributed equally. **Declaration** The author(s) declared no potential conflicts of interest with respect to the research, authorship, and/or publication of this article.

## Abstract

**Background:** To characterize C-reactive protein (CRP) changes features from patients with coronavirus disease 2019 (COVID-19) and to quantify the correlation between CRP value and clinical classification.

**Methods:** This was a bidirectional observational cohort study. All laboratory confirmed COVID-19 patients hospitalized in Xiangyang No.1 People’s Hospital were included. Patients’ general information, clinical type, CRP value and outcome were collected. Patients were grouped according to the age, clinical type and outcome, and their CRP were compared. The CRP value, age gender, and clinical type were used to build a categorical regression model to investigate the association between CRP and clinical type.

**Results:** The 131 patients aged 50.13±17.13 years old. There were 4 mild, 88 moderate, 21 severe and 18 critical cases. Statistical significance of CRP median exists between different clinical types and ages. There were 10 deaths and 121 cases have been discharged. The CRP in death group dramatically increased continuously until died, while increased firstly and decreased later in the survivor and survivor in critical type. The categorical regression model also showed that CRP and age had significant coefficient. During the first 15 days from symptom onset, the maximum of CRP ranged between 0.47-53.37 mg/L were related to mild combined with moderate type, ranged 53.84-107.08 mg/L were related to severe type, and 107.42-150.00 mg/L were related to the critical type.

**Conclusions:** CRP showed different distribution feature and existed differences in various ages, clinical types and outcomes of COVID-19 patients. The features corresponded with disease progression.

## Introduction

Coronavirus disease 2019 (COVID-19), a severe acute respiratory syndrome coronavirus 2 (SARS-CoV-2) caused deadly respiratory illness, has become a global epidemic [1-2]. The World Health Organization (WHO) declared COVID-19 as a threat to the global [3]. CRP (C-reactive protein) reflected the changes in the intensity of inflammatory response sensitively when strengthen inflammation appeared [8-9]. Researchers found CRP acted as an independent variable correlated with disease severity (severe type and critical type) and outcome through analyzed admission CRP [13] and CRP median value in severe cases was higher than critical cases (*P*<0.0001) [010]. Liu *et al*. indicated that CRP median value was significantly higher in elderly group than young and middle-aged group (*P*<0.001) [09].

Through analyzing the CRP median value in initial, progression, peak and recovery stage that were divided based on computed tomography (CT) progress, researchers found that CRP in severe type were higher than that in mild type during initial and progression stages, and CRP values were correlated with CT severity scores [18]. According to the published studies, admission CRP, maximum CRP, and mean CRP values in various stages or groups [19] were collected to detect the relationship with clinical type, outcome, lung lesion, etc. and might even be useful in the early diagnosis [20-22]. However, there was a lack of systematic research on the changes of CRP and the relationship between CRP values and the process of COVID-19 in different ages, clinical types and outcomes. In order to reveal the features of CRP in different groups and investigate the association between CRP values and clinical type, we included all confirmed COVID-19 patients hospitalized in Xiangyang No.1 People’s hospital, and the enrolled cases all discharged or died. Then analyzed all CRP results tested before admitted and during hospitalization.

## Material and methods

### Study design

This research project was a bidirectional observational cohort study. This cohort established on Feb 9, 2020, all suspected and laboratory-confirmed COVID-19 patients hospitalized in Xiangyang No.1 People’s Hospital Affiliated with Hubei University of Medicine before Feb 28, 2020 were included in this cohort. All information was traced back to the Jan 23, 2020. The last day of follow-up was on Mar 28, 2020. Admission standard and Clinical classifications were made according to the *Diagnosis Guidance for Novel Coronavirus Pneumonia* [23]. The study was approved by the ethics review board of Xiangyang No.1 People’s Hospital (No. 2020GCP012) and registered at the Chinese Clinical Trial Registry as ChiCTR2000031088. Informed consent from patients has been exempted since this study is an observational cohort study that neither involve patients’ personal privacy nor incur greater than the minimal risk.

### Data collection

Data were extracted from electronic medical records by two groups (two researchers per group) using a consistent data collection protocol and cross-checked individually. Gender, age, all CRP test results, disease onset date, outcome were collected. The data within the course of 1-30 days were statistic analyzed. The distributions of CRP median value during the course were plotted with an interval of 5 days (T1, T2, T3…Tn represented the 5-days unit successively). Two respiratory physicians classified the patients from mild to critical type and then cross-checked the results, the third expert was involved when there was disagreement. The cases were divided into 5 groups depending on their age, including: ⩽ 40, 41-50, 51-60, 61-70, and >70 years old when studied the CRP in different age groups. Patients were grouped into survivor, survivor in critical type, and death groups according to their outcomes when studied the CRP in different outcomes. Maximums of CRP during the first 15 days were collected. CRP value, age, and gender were used as independent variables (age and CRP as ordinal variables, gender as a nominal variable), clinical classification was used as dependent ordinal variables to set up the regression model.

All patients’ general information, and days from symptom onset to death, symptom onset to discharge, and symptom onset to severe/critical type were included. CRP median in various ages, different classifications, and outcomes, significance analyses between groups, CRP positive rate, changes of CRP median value over time, and categorical regression model were studied.

### CRP examination

The CRP test was conducted by the Laboratory Department of Xiangyang No.1 People’s Hospital, using Turbidimetric inhibition immune-assay. The reagent was C-reactive protein test kit (Abbott Laboratories), the test instrument is automatic biochemical immunoassay analyzer (Abbott Laboratories ARCHITECT c16200), the normal range of CRP value was 0-8 mg/L.

### Statistical analysis

All statistical analyses were performed using SPSS 20.0 Binary data were described using frequency and percentage. The normality of continuous data was checked. Mean and standard deviation were used to describe variables with normal distribution; otherwise, median (IQR) was used. Categorical data were described as frequency (%); the chi-square test was applied to assess significance between groups. Besides, the t-test and Mann-Whitney U test were separately used to compare the normal and non-normal distribution between the two groups. Categorical regression model based on optimal scaling was built for quantitative analysis. All graphs were processed using GraphPad Prism 8.0 and Photoshop CC 14.2 software.

## Results

This study included all of the suspected and laboratory-confirmed 542 patients till on 28^th^ Feb 2020. Among the 542 cases, the nucleic acid tests in 142 cases were positive but 9 cases that have data stored in other hospitals cannot be traced and 2 were infants. Therefore, 131 cases were included in the study.

### General information

Among the 131 included cases (63 M/ 68 F), 123 cases of which were discharged from the hospital and 10 cases died. The average age was 50.13±17.13 years old. The time from symptom onset to severe type was 8.15±5.29 days, symptom onset to critical type was 11.83±6.01 days, symptom onset to death was 18.30±9.65 days, and symptom onset to discharge from the hospital was 26.87±9.19 days.

### CRP results

The 131 cases underwent 37 laboratory indicators contained 24052 tests in outpatient and hospitalization. This study included all of the CPP results 724 times of tests totally, account for 3.01% of the all results of the indicators.

### The features and differences of CRP in different ages

The 41 cases in ⩽40 years old group underwent 163 tests (1-36 days) of CRP, and the median was 3.62(1.43-11.28) mg/L, among which 30.67% of the results exceed the upper limit normal value (ULN). 159 tests during the first 30 days with a median of 3.62(1.43-12.49) mg/L, and 30.82% tests were higher than the normal extent; The 28 cases in 41-50 years old group underwent 143 tests (1-51 days) with the median of 7.35(1.89-20.60) mg/L. Among them, 46.85% tests exceed the ULN. 124 tests underwent during the first 30 days with a median value of 8.08(1.87-24.25) mg/L, and 50.00% results were over the normal range; The 23 cases in 51-60 years old group underwent 101 tests (1-39 days), and the median was 5.38(2.52-24.58) mg/L. Above them, 44.55% tests surpassed the ULN. 92 tests underwent during the first 30 days with the median as 6.78(2.83-26.14) mg/L. And 48.91% of the results were higher than the ULN; The 21 cases in 61-70 years old group underwent 176 tests (1-39 days), and the median was 13.54(4.90-50.82) mg/L, among them 64.04% tests exceed the ULN. 153 tests underwent during the first 30 days with the median value as 16.28(5.42-53.55) mg/L, and 67.32% tests were higher than the normal extent; The 18 cases in >70 years old group underwent 141 tests (1-37 days), and the median was 35.26(12.19-92.13) mg/L, 78.01% tests were higher than the ULN. 133 tests underwent during the first 30 days, with the median as 37.72(13.90-97.60) mg/L. 81.95% of the results exceed the normal extent. The CRP median and the corresponding changes in different age groups were shown in Table 1 and Figure 1. According to the figure, the peak value (84.49 mg/L) in T3 and the abnormal interval was T1-T6 in >70 years old group, which was different from other groups.

**Table 1.**
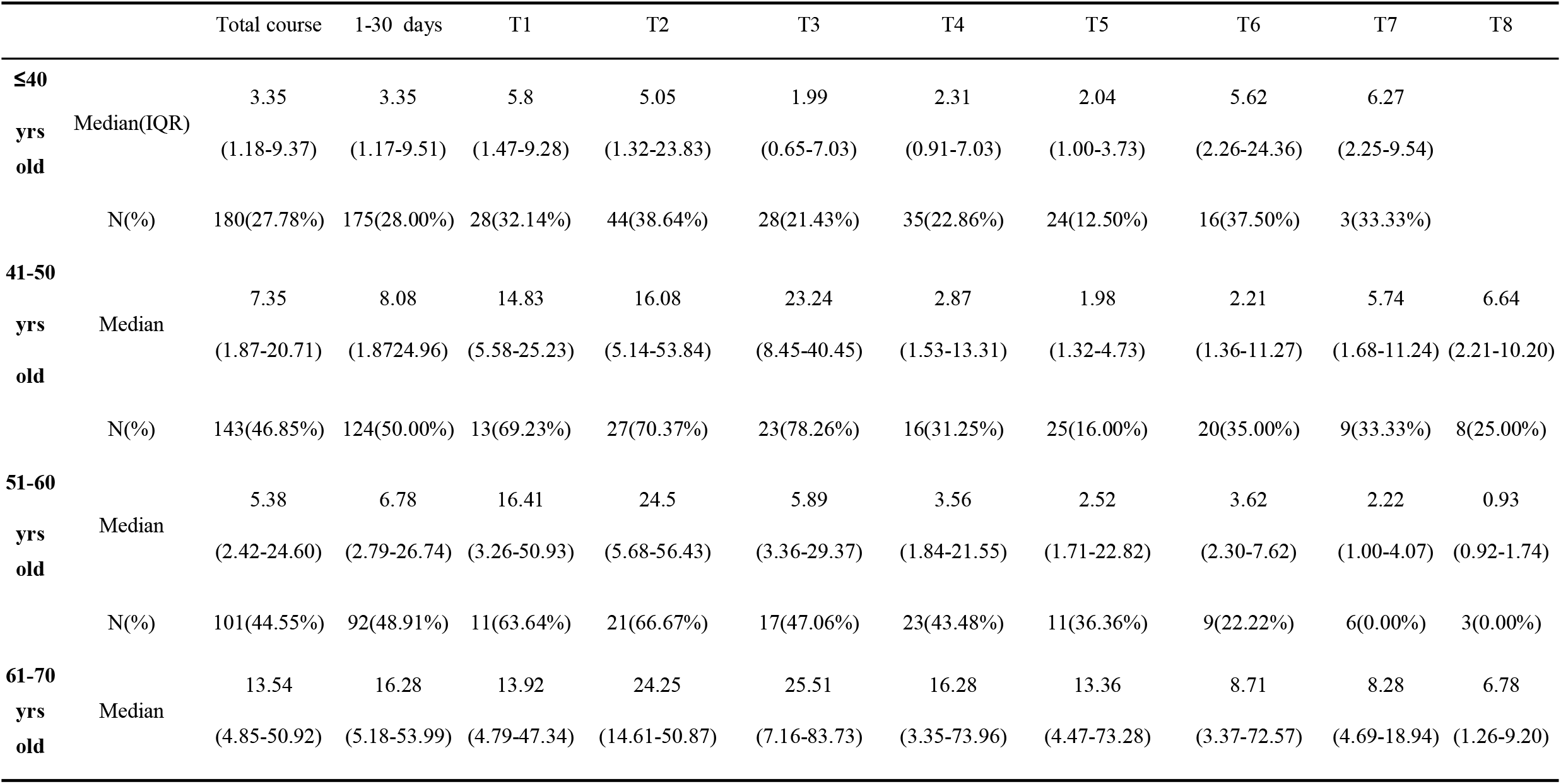
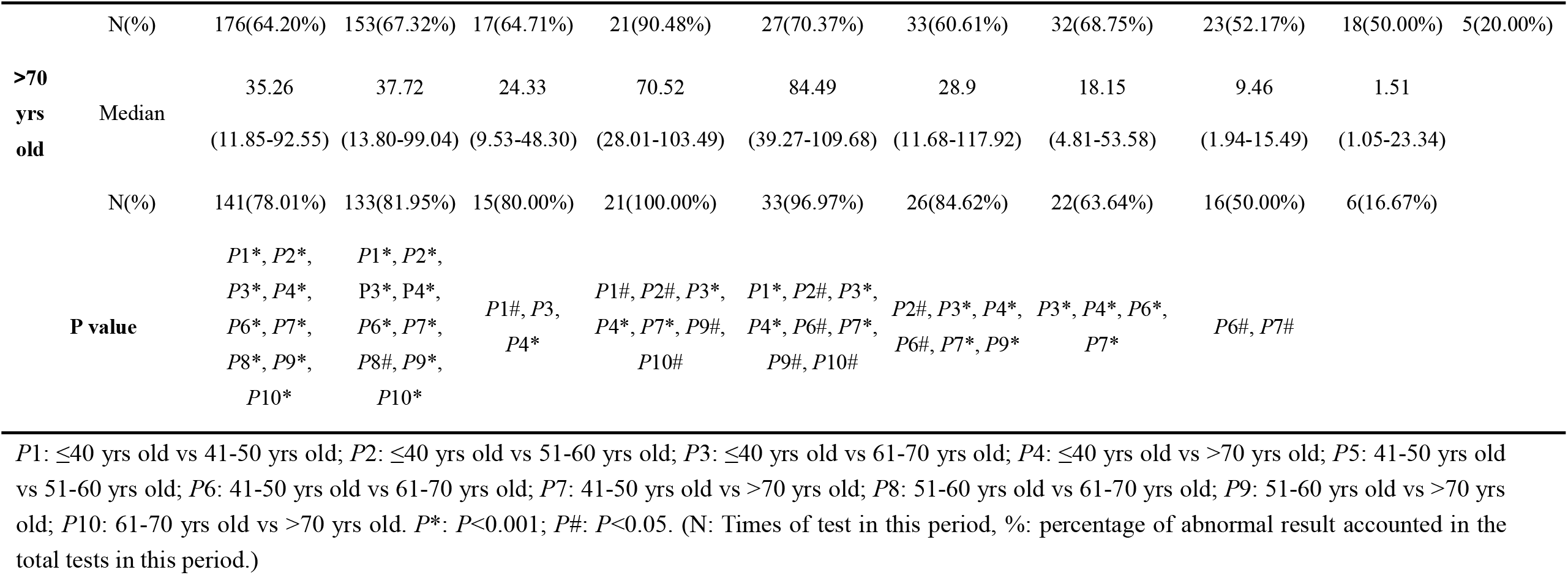
The features of CRP in the COVID-19 patients with different age groups

**Figure 1.**
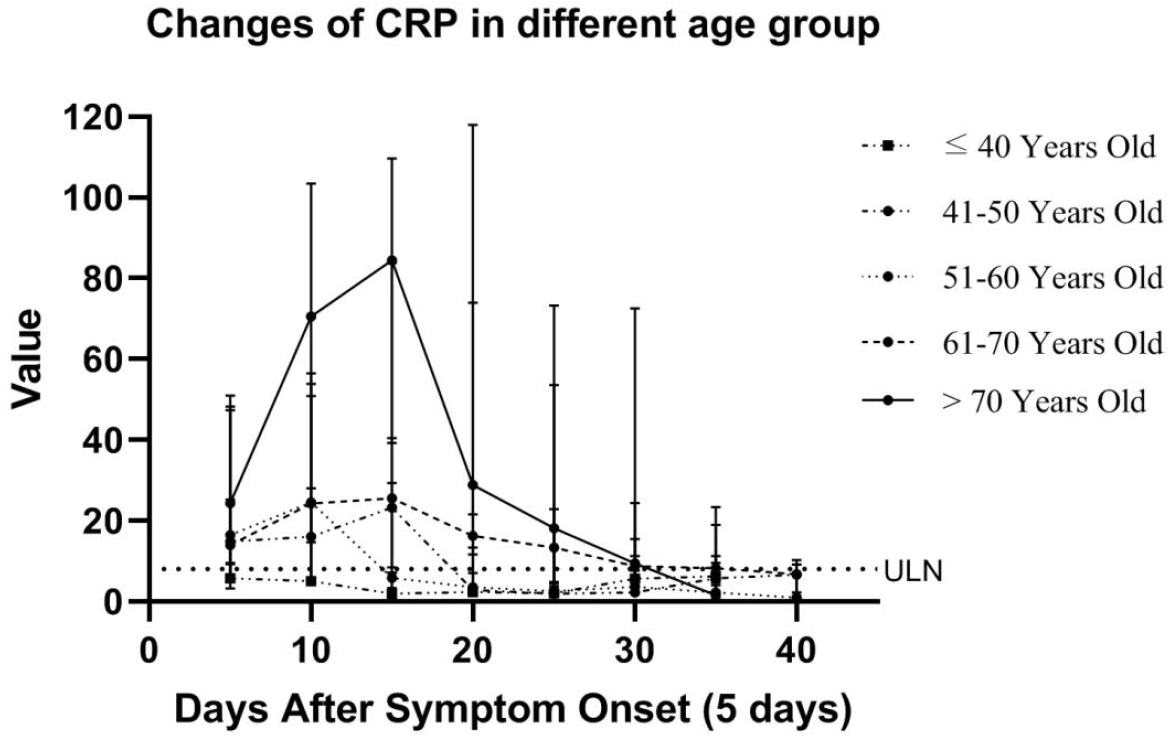
The distribution over time of CRP median in the overall and the different age groups

Differences within the first 30 days between different age groups (*P*<0.05), CRP median value in T1 and T3 were significantly different between ⩽40 and 41-50 years old group (*P*<0.05); in T2-T3 were significantly different between ⩽40 and 51-60 years old group; in T2-T5 were significantly different between ⩽40 and 61-70 years old group (*P*<0.05) and also ⩽ 40; in TI-T5 were significantly different between ⩽ 40 and >70 years old group (P<0.05); in T4-T6 were statistically different between 41-50 and 61-70 years old group (*P*<0.05); in T2-T5 were statistically different between 41-50 and >70 years old group (*P*<0.05); in T2-T4 were significantly different between 51-60 and >70 years old group; in T2-T3 were significantly different between 61-70 and >70 years old group (*P*<0.05) (Table 1).

### The features and differences of CRP in different clinical types

The 4 mild cases aged 41.25±23.77 years old underwent 21 tests (1-26 days) of CRP and the median was 1.99(1.09-3.21) mg/L. One of the test exceeded the normal range; The 88 moderate cases aged 45.26±14.73 years old underwent 350 tests (1-51 days) with the median as 4.72(1.79-15.65) mg/L.37.14% of the tests exceeded the normal range. And 331 tests underwent during the first 30 days with the median as 4.93(1.80-17.10) mg/L, of which 38.37% tests were higher than the normal extent; The 21 severe cases aged 56.05±15.30 years old underwent 168 times of CRP tests (1-45 days), the median was 11.81(2.58-28.30) mg/L. There were 55.95% tests exceeded normal range. The 141 tests underwent during the first 30 days, and the median was 13.93(4.15-31.55) mg/L, of which 62.41% of the tests were higher than the normal extent; The 18 critical cases aged 69.00±14.08 years old underwent 184 tests (1-38 days), the median was 58.99(15.27-107.48) mg/L. The 86.96% of the tests had exceeded normal range. 168 tests underwent during the first 30 days, and the median was 68.12(18.27-110.27) mg/L, of which 90.48% tests were higher than the normal extent. The CRP median and the corresponding changes in different clinical types were shown in Table 2, Figure 2. According to the changes, median value in critical type showed a peak value (89.09 mg/L) in T3, and the abnormal interval was T1-T6, which was different from other classifications.

**Table 2.** The features of CRP in the COVID-19 patients with different clinical types

| Period | Mild type |  | Moderate type |  | Severe type |  | Critical type |  | P value |
| --- | --- | --- | --- | --- | --- | --- | --- | --- | --- |
|  | Median(IQR) | N(%) | Median(IQR) | N(%) | Median(IQR) | N(%) | Median(IQR) | N(%) |  |
| Total course | 1.99(1.09-3.2) | 21(4.76%) | 4.65(1.79-15.36) | 352(36.93%) | 11.84(2.58-28.30) | 168(55.95%) | 48.8(10.91-104.53) | 199(80.40%) | *P1\**, *P2\**, *P3\**, *P4\**, *P5\**, *P6\** |
| 1-30 days |  |  | 4.75(1.79-16.98) | 333(38.14%) | 13.93(4.15-31.55) | 141(62.41%) | 56.57(14.39-107.34) | 152(83.52%) | *P4\**, *P5\**, *P6\** |
| T1 | 0.71(0.64-1.2) | 5(0.00%) | 8.58(3.89-20.77) | 52(53.85%) | 13.92(5.57-62.57) | 13(61.54%) | 26.12(15.78-45.14) | 13(92.31%) | *P1#*, *P2#*, *P3\**, *P5#* |
| T2 | 2.50(1.44-4.5) | 6(16.67%) | 12.79(3.76-28.93) | 75(56.00%) | 24.25(17.38-39.22) | 23(95.65%) | 71.68(37.73-101.89) | 30(83.33%) | *P1#*, *P2\**, *P3#*, *P4#*, *P5\**, *P6#* |
| T3 | 2.94(2.59-3.4) | 6(0.00%) | 9.21(2.80-22.36) | 62(51.61%) | 32.04(18.81-39.77) | 22(77.27%) | 88.29(47.69-111.21) | 38(89.47%) | *P1#*, *P2\**, *P3\**, *P4#*, *P5\**, *P6\** |
| T4 | 2.03(1.9-2.2) | 2(0.00%) | 2.61(1.51-4.96) | 66(22.73%) | 14.10(3.13-25.62) | 28(67.86%) | 68.88(13.9-120.24) | 37(83.78%) | *P3#*, *P4\**, *P5\**, *P6\** |
| T5 | 1.34(1.08-1.6) | 2(0.00%) | 1.92(1.27-3.89) | 47(10.64%) | 5.58(2.43-28.35) | 31(45.16%) | 31.03(15.17-115.00) | 35(80.00%) | *P4\**, *P5\**, *P6\** |
| T6 |  |  | 2.94(1.59-4.73) | 31(16.13%) | 5.71(2.23-11.47) | 23(34.78%) | 14.5(8.75-120.12) | 29(75.86%) | *P4#*, *P5\**, *P6\** |
| T7 |  |  | 4.33(2.95-7.44) | 11(27.27%) | 1.8(1.07-6.87) | 19(21.05%) | 9.23(7.29-29.40) | 12(58.33%) | *P5#*, *P6\** |
| T8 |  |  | 3.78(1.24-7.01) | 6(0.00%) | 0.97(0.93-1.74) | 9(22.22%) | 7.10(6.04-8.30) | 4(25.00%) |  |
| T9 |  |  | 2.86(2.71-3.01) | 2(0.00%) |  |  | 1.3 | 1(0.00%) |  |

Median (IQR): median value (interquartile range); N: Times of test in this period; %: percentage of abnormal result accounted in the total tests in this period; *P1*: Mild type vs moderate type; *P2*: Mild type vs severe type; *P3*: Mild type vs critical type; *P4*: Moderate type vs severe type; *P5*: Moderate type vs critical type; *P6*: Severe vs critical type. *P\**: $P<0.001$ ; *P#*: $P<0.05$ . (N: Number of CRP tests, %: the abnormal rate)

**Figure 2.**
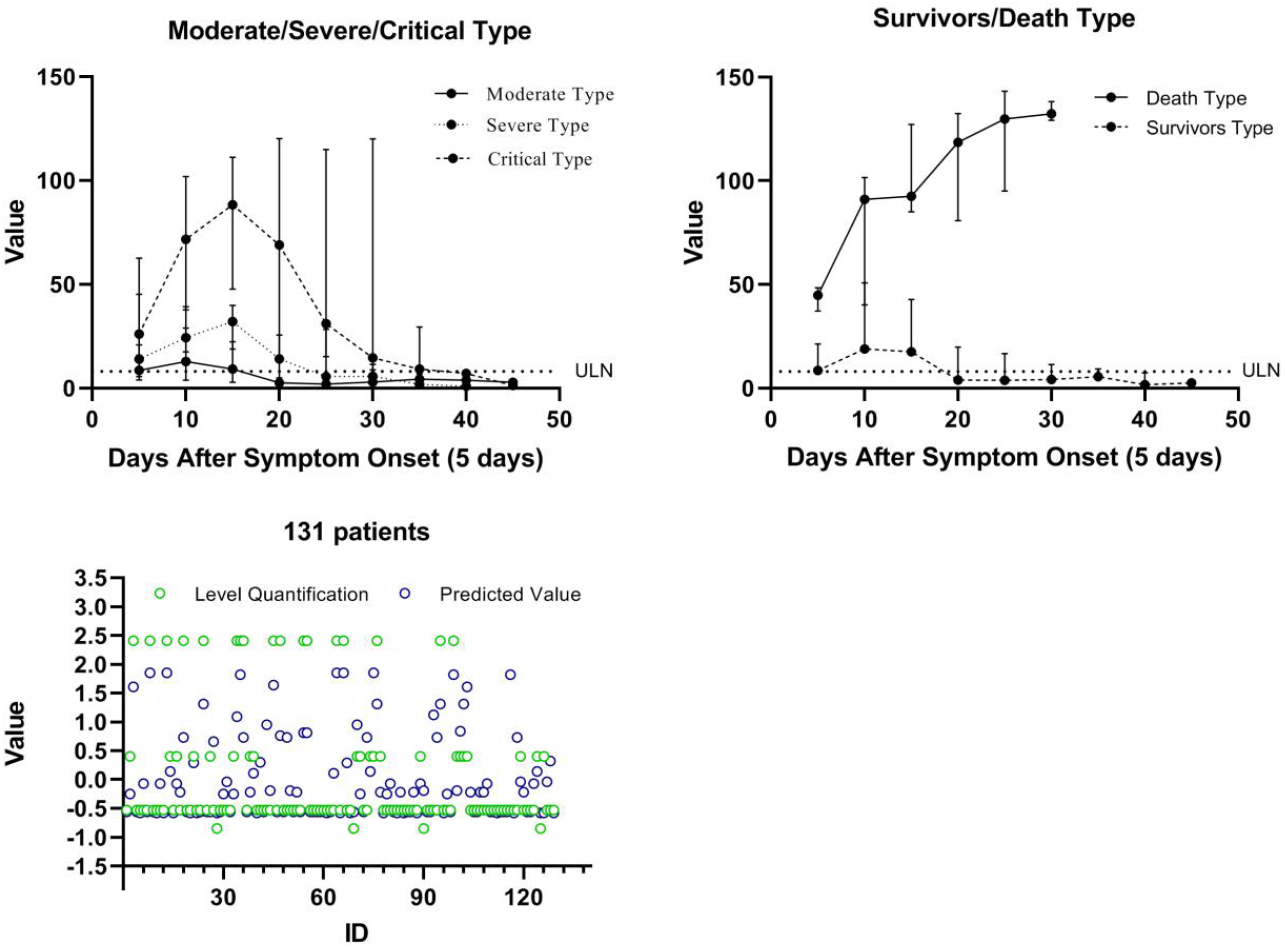
The distribution over time of CRP median in different clinical type and outcome and scatterplot of severity level quantification and categorical regression model’s output quantification values

There was a statistically significant difference (*P*<0.05) among mild, moderate, severe, and critical type. In T1, moderate, severe, and critical type all had CRP under 30.00 mg/L, but after T1, moderate and severe type had a slight increase in CRP, while critical type had a sharp increase in CRP. Significant differences (*P*<0.05) can be found between moderate and severe type in T2-T6, between moderate and critical type in T1-T6 (*P*<0.05) and also between severe and critical type in T2-T6 (*P*<0.05) (Table 2).

### The features and differences of CRP in different outcomes

Among the 131 cases, there were 10 cases died; the remaining 121 cases have been discharged. The 121 survivors underwent 655 tests and the median was 7.40(2.29-24.60) mg/L. 48.24% of the tests exceeded normal range; The 10 death cases underwent 69 tests of CRP with the median as 101.73(60.89-129.43) mg/L and 100.00% of the tests exceeded normal range. The CRP median and the corresponding changes in different outcomes were shown in Table 3, Figure 2. Death group showed a increasing trend all along even when the other groups decreased after T3, and the abnormal interval was T1-T6, which was different from survivor group.

**Table 3.**
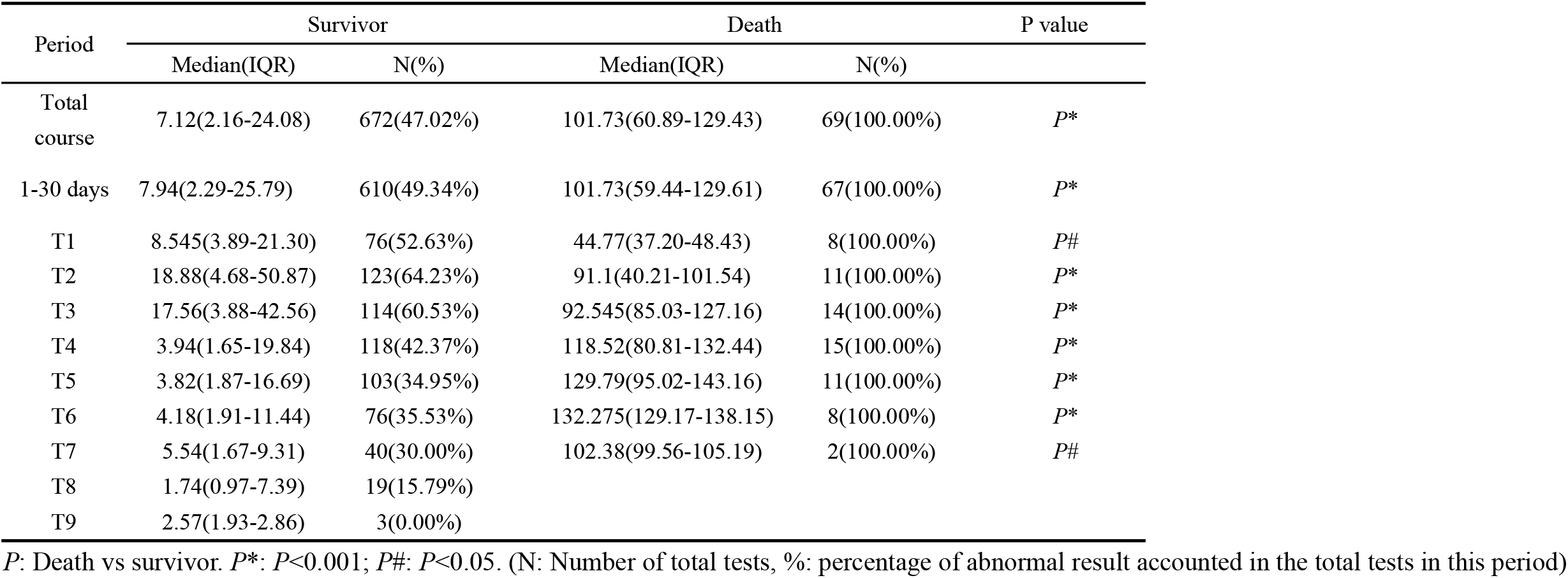
The features of CRP in the COVID-19 patients with different outcomes

Statistically differences could be found between death cases and survivor cases. In T1-T6, statistically significant difference (*P*<0.05) could be found between survivor cases and death cases (Table 3).

### Quantify the correlation between CRP and clinical types base on categorical regression

The maximum CRP in the first 15 days after symptom onset was collected to build the categorical regression based on optimal scale. The model output showed age (*P*<0.01) and CRP (*P*<0.001) had significant coefficients while gender (*P*>0.05) did not have significant coefficient. The quantification of CRP categories showed CRP ranged between 0.47-53.37 mg/L was related to mild combined with moderate type. CRP ranged 53.84-107.08 mg/L was related to severe type, and 107.42-150.00 mg/L was related to critical type. The quantification of age showed 15-61 years old was related to mild combined with moderate type, 62-70 years old was related to the severe type, and 71-90 was related to critical type. The model expression was Q-level =0.553*Q_CRP+0.014*Q_sex+0.309*Q_age. The comparison between output severity quantification of different clinical type output by this model and the real severity quantification of the clinical type was shown in Figure 2. The model cannot distinguish mild and moderate type, so we combined the 2 types and the accuracy rate of such output on three types (mild combined with moderate, severe, critical) was 75.97%. Moreover, if severe and critical type were combined as well, the accuracy rate was 82.17% to discriminate the two combined type (Table 4, Figure 2).

**Table 4.**
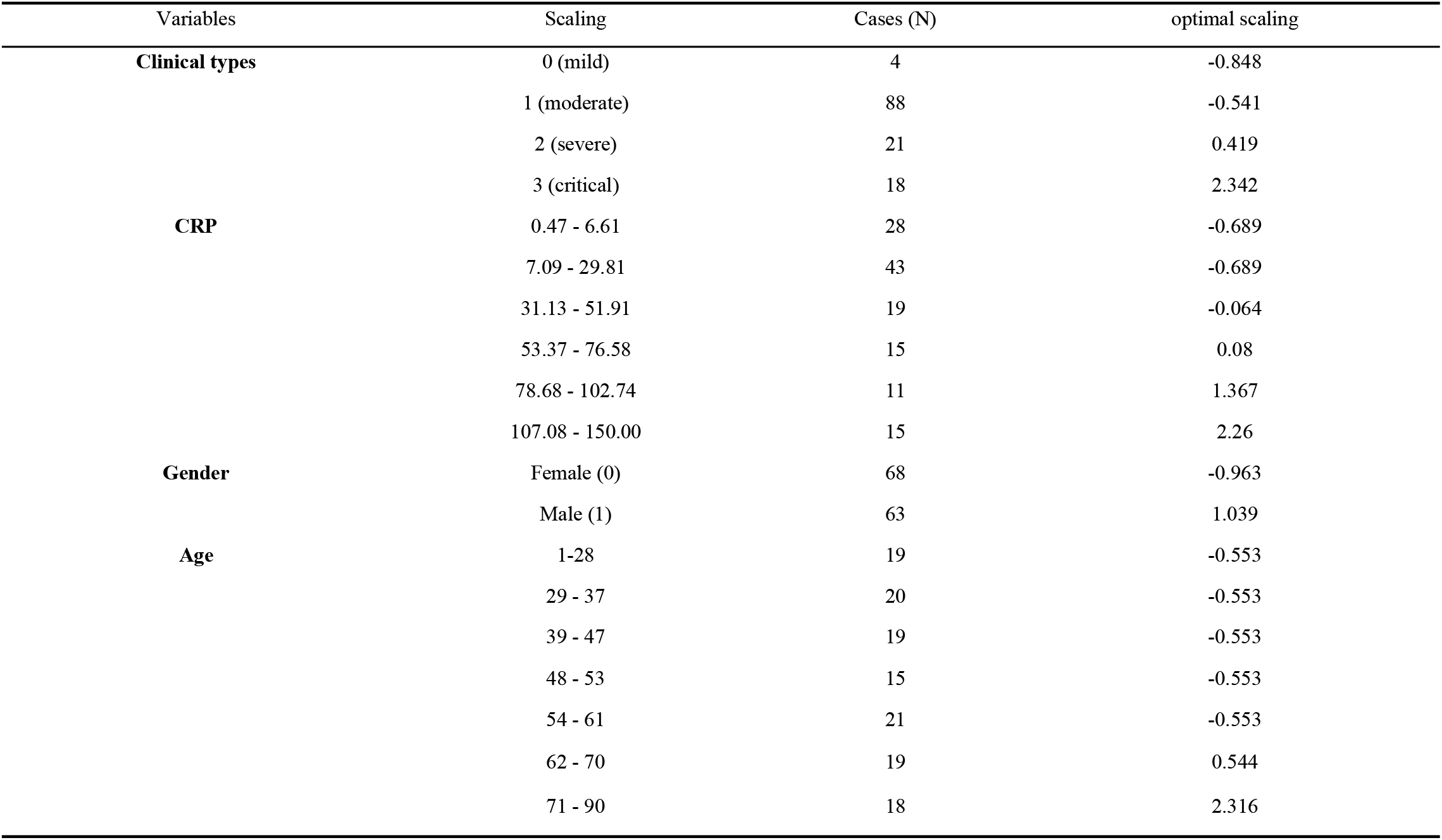
Variable categories and quantifications based on optimal scaling

## Discussion

We enrolled hospitalized 131 SARS-CoV-2 test positive cases. To avoid the influence of patients with long hospitalization period, we statistically analyzed the data within 1-30 days according to 26.87±91.9 days the time from symptom onset to discharge from hospital.

Most of the patients in this cohort were >40 years old, and older age reflected much more serious severity according to the clinical classification. Meanwhile, CRP in different clinical types showed higher CRP related to severer disease, suggesting that differences in CRP distribution might exist within various ages. In our study, the CRP result of patients ⩽40 years old all sustained in a normal extent. Patients aged 41-50 and 51-60 years old separately had an abnormal period during T1-T3 and T1-T2, the peak value were 23.24 mg/L and 24.5 mg/L, respectively; Patients aged 61-70 and >70 years old both had a prolonged abnormal period of T1-T6 and increased peak values. Among them, the peak value in >70 years old group was increased up to 84.49 mg/L. In summary, the results revealed that older age might induce higher median and peak value, and longer abnormal period except for the 51-60 years old group, which indicated older age induced more severe inflammatory response, more serious illness, and a higher rate of developing into severe and critical types. But the reason age influenced the inflammatory response and the intensity need further study.

The abnormal rate in mild, moderate, severe and critical type was 0, 37.14%, 55.95%, 86.96%; median was 1.99 mg/L, 4.72 mg/L, 11.84 mg/L, 58.99 mg/L; peak value was 2.94 mg/L (T3), 12.79 mg/L (T2), 32.04 mg/L (T3), 89.09 mg/L (T3), respectively. The median in the total course and different periods in mild type kept in normal extent, which suggested gently tissue injury or inflammation in this type. Median values in the total course were normal but increased in T1-T2, and the peak value (12.79 mg/L) was situated in T2 in moderate type, inflammatory response mainly appeared within the first 10 days. Median values of severe and critical types kept increasing during TI-T3. The peak value was 32.04 mg/L and 89.09 mg/L in T3 and severe type recovered in T5, indicating serious tissue injury with severe inflammatory response in these two types. There were significant differences of CRP in different severity cases. The more severity of the clinical classification associated with the higher median, peak value, incidence of abnormal values, the faster-risen speed, the longer outlier interval, and the slower declined speed of CRP.

In moderate type, the CPR median peak value located in T2, and CRP showed persistently decreased after T2. Further increased CRP after 10 days indicated the great risk to develop into severe and critical type because the peak values of CRP in severe and critical type were situated in T3. The upper limit value within T2 in moderate and severe type was 28.93 mg/L and 39.22 mg/L respectively, the median value in critical type was 87.09 mg/L. These values suggested that patients might transform into severe or critical illness when CRP>39.22, especially >87.09 mg/L. In T3, the upper limit value was 4.96 mg/L, 39.77 mg/L in moderate and severe type, respectively, and the median was 89.09 mg/L in critical illness. Patients might develop into severe and critical when CRP >39.77 mg/L, especially >89.09 mg/L. This also suggested that CRP differences appeared during T2-T3 (5-15 days) in different clinical types, which was promising in the prognosis and forewarning.

Cases with different outcomes have significant differences in features of CRP. The median CRP of survivor and death groups were 7.40 mg/L, 101.73 mg/L, the positive rate was 48.24%, 100.00%, the peak value was 18.03 mg/L, 132.28 mg/L, and abnormal interval were T1-T3, T1-T6, respectively. Death cases have statistically significant differences from the survivors. The higher CRP indicated a higher risk of death. CRP>44.77 mg/L (the median value in death) in T1 might prompt high death. In the T2 period, survivor and death group existed a significant difference. It has a high risk of death when CRP>91.10 mg/L (the median value in death); In T3 period, the risk of death was high when CRP>92.55 mg/L (the median value in death). The risk of developing into critical type or even death increased if CRP elevated after T2. The risk of death was high when it progressively increased after T3. This finding was useful for the diagnosis and prognosis, especially in the early stage.

Different from the survivors with the changes gradually decreased to a normal level, CRP rapidly elevated within 10 days and then increased persistently but slowly after 10 days until died in the death group. In the early stage, virus infection induced CRP to increase because of injury and inflammatory response. In the advanced-stage, virus infection might also combine bacterial infection; thus, CRP maintained at a high level and resulted in death due to single or multi-organ failure. In summary, the more serious disease showed a higher median and peak value, the longer elevating period of disease course, the faster speed of increasing, and the slower restoration.

The time from symptom onset to developing into critical type was 11.83±6.01 days; thus, we selected a maximum of CRP in the first 15 days was collected to establish classification model. The result showed that the maximum of CRP during the 15 days ranged 0.47-53.37 mg/L, 53.84-107.08 mg/L, 107.42-150.00 mg/L was related to mild combined with moderate type and severe type, critical type, respectively. However, there were only 4 mild cases; thus, the model couldn’t differentiate mild type exactly. According to the clinical type regression model, classification on mild combined with moderate, severe, and critical type has an accuracy rate of 75.97% and might influenced by the overlapped data in severe and critical type. Since our aim of exploring the relationship between CRP and severity level was focused on discriminating severe and critical types. Therefore, we combined severe and critical type basing on output results to discriminate the 2 combined classifications, and the accuracy rate was 82.17%. Such findings can be regarded as standards for severity classification in this cohort and CRP can be used to distinguish clinical type if the output of classification verified by other data.

The cases in this cohort were obtained from single hospital’s inpatient cases, and the evidence value was limited thus it was impractical to analyze daily CRP data or describe its daily change. We didn’t analyze the data after discharge from hospital. Multi-centered and more samples are needed to verify the reference standard of CRP in severity classification and prognosis.

In conclusion, The CRP median value of overall tests and most of the results were abnormal with the manifestations of increasing during the early to middle stage. Older age, more aggravated illness, and worse outcomes always presented as higher positive rate, median value, peak value, and longer abnormal CRP duration. CRP value ranged between 0.47-53.37 mg/L was related to mild combined with moderate type, 53.84-107.08 mg/L was related to severe type, 107.42-150.00 mg/L was related to the critical type. The changes in CRP could reflect the severity of COVID-19. Furthermore, analysis and discrimination of classification and prognosis in COVID-19 might be applied if verified through other data.

## Data Availability

Anyone who wishes to obtain the original data of this study with reasonable purposes can contact the correspondent author via email.

